# Effects of on Low T3 syndrome astrocyte homeostasis in patients with bisphenol A-associated intracerebral hemorrhage

**DOI:** 10.1101/2023.06.12.23291309

**Authors:** Guoping Wang, Jinhua Chen, Zhongdong Qiao, Dongxing Guo, Ping Guo, Aiven Wang, Wanli Sun, Jiyuan Lyu

**Affiliations:** Affiliated Changzhi People’s Hospital of Shanxi Medical University; Shanghai Jiao Tong University.; Shanxi Medical University Basic medical college.; Changzhi Medical College Peace Hospital.; First Hospital of Shanxi Medical University.

## Abstract

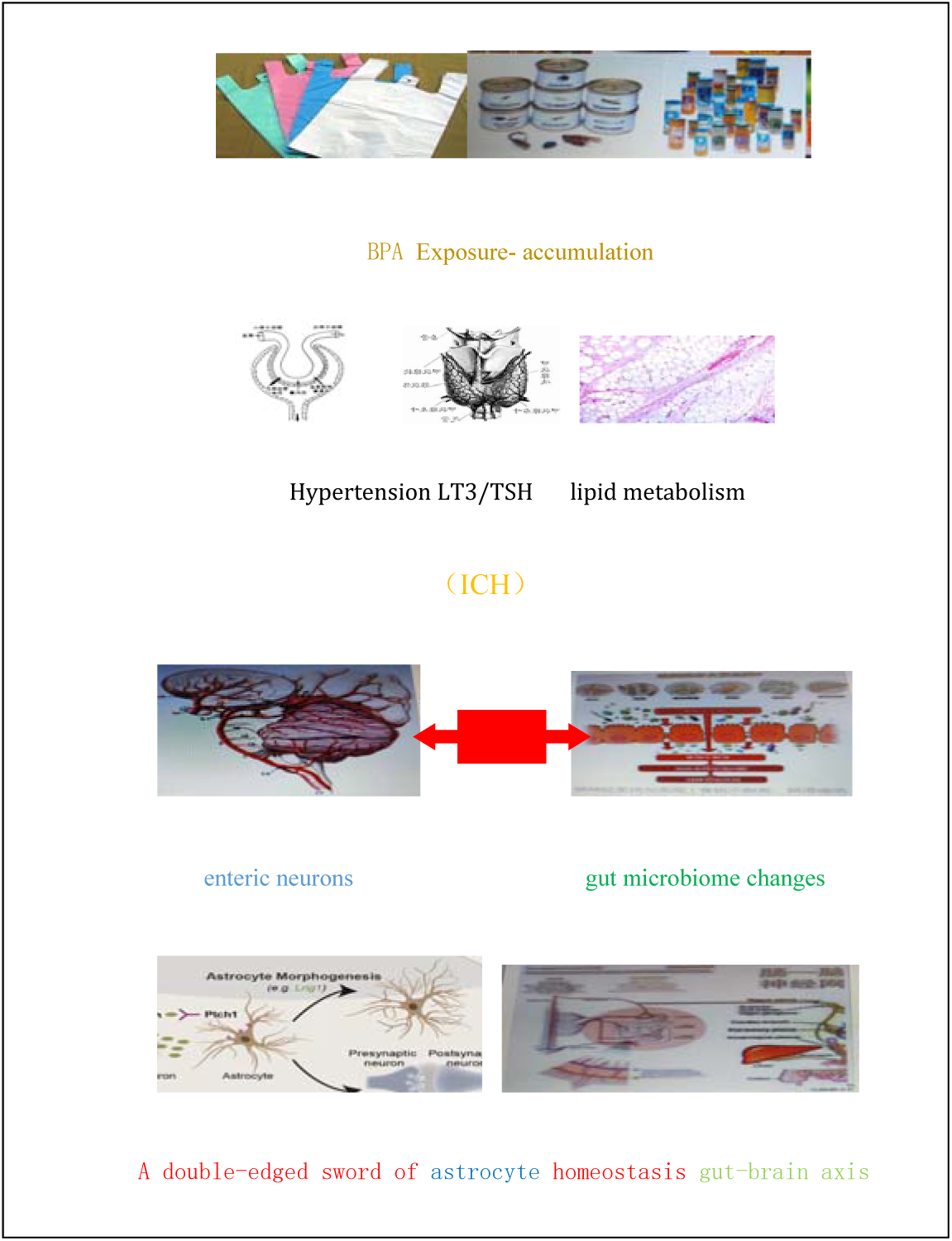

**Background and Purpose:** We aimed to assess sex differences in lipid metabolism disorders in patients with BPA-related intracerebral hemorrhage, as well as changes in β3-adrenoceptors and oxidative stress parameters, and to explore potential mechanisms of intestinal flora on the functional homeostasis of cerebral astrocytes.

**Methods:** Here, a multicenter longitudinal retrospective study was conducted in 200 patients with intracerebral hemorrhage compared with normal controls. Serum thyroid function, blood lipids and subclasses, superoxide dismutase and malondialdehyde were detected. Active oxygen species and reactive oxidative products in brain tissue were detected by fluorescein labeling apparatus. Lipid metabolism and oxidative stress regulate messenger proteins, as well as changes in intestinal flora and astrocyte expression.

**Results:** Serum lipoprotein β levels of LT3 male patients were significantly increased, with gender differences compared with female patients. β3-adrenergic receptor and Neuregulin –1 expression changes, targeted regulation of fat and reduce oxidative stress response. The increased expression of messenger protein prompts activation of brain astrocytes. The intestinal flora study found that the alpha diversity index in the low T3 group was higher than that in the control group, and Spearman correlation analysis showed that low T3 level was negatively correlated with lipid metabolism disorders and oxidative stress.

**Conclusions:** Bisphenol A and oxidative stress were significantly related to the high risk of cerebral hemorrhage. The changes of inflammatory cytokines and the ratio expression of connexin 43/Yes associated proteins in the gut brain axis drive activated astrocytes and microglia to jointly maintain the homeostasis of cerebral vascular microenvironment.

*Trial registration:* The trial is registered at clinical.gov (www.medresman.ChiCTR1900023626)

## Introduction

Intracerebral hemorrhage (ICH) accounts for 10%-15% of stroke cases, and the assessment of ICH risk factors and the study of toxicological mechanisms are clinically challenging ^1–3^. Insight into the downregulation of connexin 43 (Cx43) in activated astrocytes associated peripheral genotoxicity, immune cell dynamics and stroke risk analysis ^4, 5^. To this end, it may be strategically positioned to explore the unique field of interaction between glial cell activation, self-protective nerve cell function, immune cell inflammatory response and long-term prognosis of ICH ^6^. Focus on changes in biotoxicity, neuro-cytotoxicity and inflammatory markers. In order to further evaluate the relationship between lipid metabolism and intestinal flora after ICH, it is helpful to provide a unique and deep new insight into the molecular study of brain microenvironment changes in ICH patients.

Bisphenol A(BPA) has estrogen-like antiandrogenic effects, and is associated with changes in thyroid function, lipid metabolism, and intestinal flora, as well as increased levels of tissue cell oxidative stress (OS) response, inducing overproduction of reactive oxygen species (ROS) ^7–9^. Promote immune inflammatory cytokines activity and neurotoxicity, can cause cerebrovascular injury. β-adrenergic signaling modulates adipocyte anabolic processes, while hypo-triiodothyronine amplifies cyclic adenosine phosphate (cAMP) messengers, leading to lipid metabolism disturbances ^10^. Neuregulin 1 (NRG1) has immunoreactive changes in the number of intestinal neurons that respond to weakened OS and have neuro-cell protective effect^11^. Lipo-toxicity and neuro-cytotoxicity interact with multiple organ functional mechanisms, possibly involving receptor pathways, immune system function, inflammatory responses, and epigenetic mechanisms ^8, 12^. Intestinal microorganisms can exert influence on the immune system and cerebrovascular through neurotransmitters and metabolites. In particular, the gut-brain axis is involved in the homeostasis of the central nervous system through the regulation of neuro-astrocytes ^13^.

## Methods

The data that support the findings of this study are available from the corresponding author upon reasonable request.

### Patients

Clinical data of ICH patients admitted to a number of hospitals from February 2014 to April 2018 were collected through systematic sampling, multi-center longitudinal and randomized case-control studies. 200 patients with ICH ranged in age from 18 to 51 years, with an average age of 38±8.3 years. They were divided into two groups according to sex: female (FMG, n=90 cases) and male (MG, n=110 cases). Meets standards of the American Stroke Association, the Cardiovascular and Stroke Care Council, and the Board of Clinical Neurology ^15^. Obesity diagnosis: According to the obesity diagnosis criteria formulated by WHO for the Asia-Pacific region: normal weight group (MIB < 25kg/m2) and obesity group (MIB > 25kg/m2) ^16^. Hypertension diagnosis criteria: systolic blood pressure ≥140 and diastolic blood pressure ≥90mmHg (1mmHg = 0.133 kpa) ^17^. 24 hours ambulatory blood pressure (24hABP) and follow-up criteria are seen (supply data online). Exclusion criteria: (1) moderate or above impairment of liver and kidney function, visceral malignancies, history of diabetes, family history of hypertension, and previous aspirin use. (2) Familial hypercholesterolemia (3) cerebrovascular malformations and cerebrovascular tumors, cerebrovascular angiography. Normal health controls were required to undergo a physical examination at a hospital health examination center and, according to the criteria, strictly select healthy individuals (n =200) matched for age and sex.

Full descriptions of other materials and methods are given in the data supplement.

### Statistical analysis

General demographic data are continuous variable mean ± SD. Comparison of qualitative data: one-way analysis of variance and Chi-square test, normality test of indicators. The μ-test was used to compare the mean values of two large samples. F test was used to compare the three groups of data. Linear correlation analysis was performed for serum BPA concentration, biomarkers and gap proteins, and regression analysis was used to determine the correlation between BPA concentration and thyroid function. Logistic regression model was used to analyze the combination and cross effects of serum BPA, CX43, YAP and intestinal flora expression levels, and to calculate the relative hazard ratio attribution ratio and interaction index. SPSS v20.0 software (SPSS Inc, USA) was used for statistical analysis, and the difference was statistically significant (P<0.05).

### Ethical approval

All participants in this study signed informed consent. This protocol was approved by the Ethics Committee of Affiliated Changzhi People’s Hospital of Shanxi Medical University (No: 2013093).

## Results

There were 109 patients in LT3 group, 80 (73%) in MG group, and 29 (26%) in FMG group. LT3 subgroup, LT3-A (n=61), LT3-b (n= 26). 91 patients in SCH group had FMG 60 (66%) and MG 31 (34%). Criteria for abnormal thyroid function: FT3≤1.5pg/ml in LT3 group, FT3 0.9-1.5pg/ml in LT3-A group, and FT3 0.3-0.8pg/ml in LT3-B group. SCH group TSH≥10μ U/ml.

FIG. 1. A: Compared with the control group, BPA levels in LT3 and SCH groups were significantly higher, and MG levels were significantly higher than FMG levels, with gender differences. B: Compared with the control group, Apo-β, apo CIII, Apo-β /apoA1 ratio BMI, HOMA-IR, TG were significantly increased in LT3 and SCH groups, while HDL-C was significantly decreased. However, compared with FMG group, Apo-β and apo CIII in MG patients showed an increasing trend in both LT3 and SCH groups, and only LT3-MG group showed a statistically significant increase, indicating a significant gender difference in lipid metabolism. C: Compared with the control group, FT3 in LT3 group was significantly decreased, while TSH in SCH group was significantly increased; The proportion of ICH-LT3 in males was greater than that in females. In the ICH-SCH group, females accounted for more than females, indicating that different stress thyroid function states in ICH presented different gender ratios. D: Compared with the control group, the serum levels of 3-nitrotyrosine and malondialdehyde (MDA) in ICH patients were significantly increased, while superoxide dismutase (SOD) was significantly decreased. Compared with LT3-FMG, the increase of 3-nitrotyrosine was more significant in LT3-MG patients, indicating that oxidative stress products were also gender-specific. E: Compared with the control group, DBP and 24hDBP in ICH patients were significantly increased. F: Compared with the control group, IL-1β, TRL-4, NF-kBP65 and MYD88 in ICH patients were significantly increased, with statistical significance (P < 0.05).

**Table 1.**
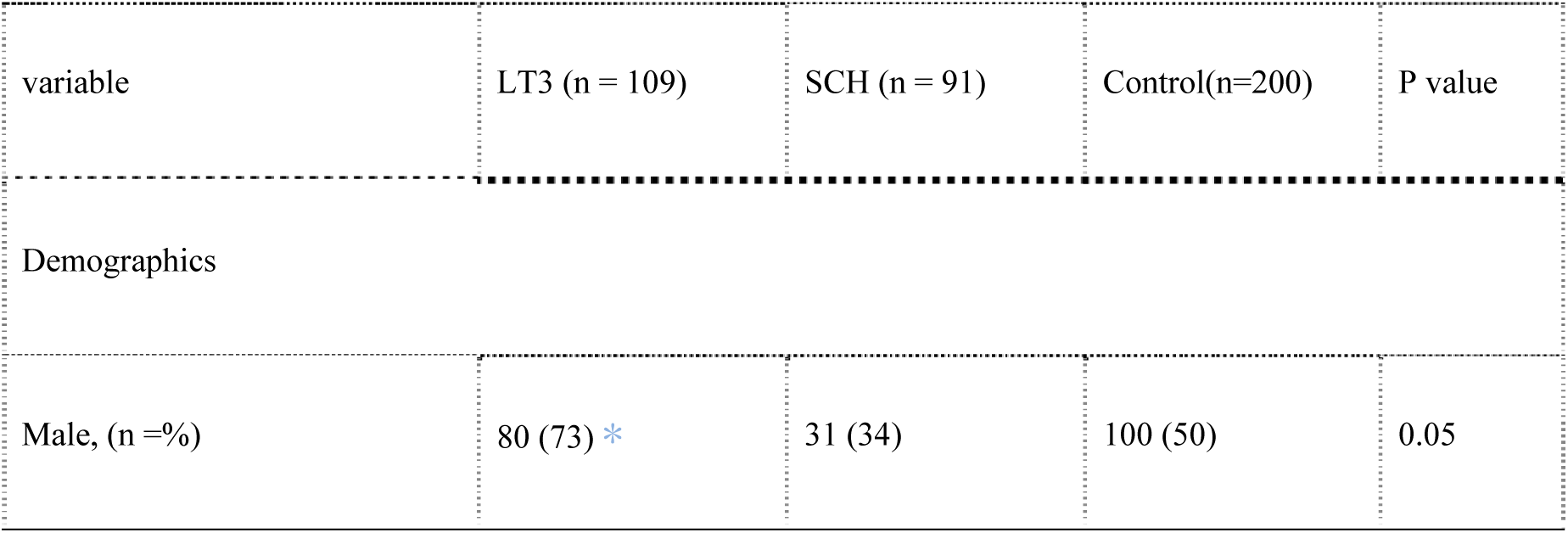
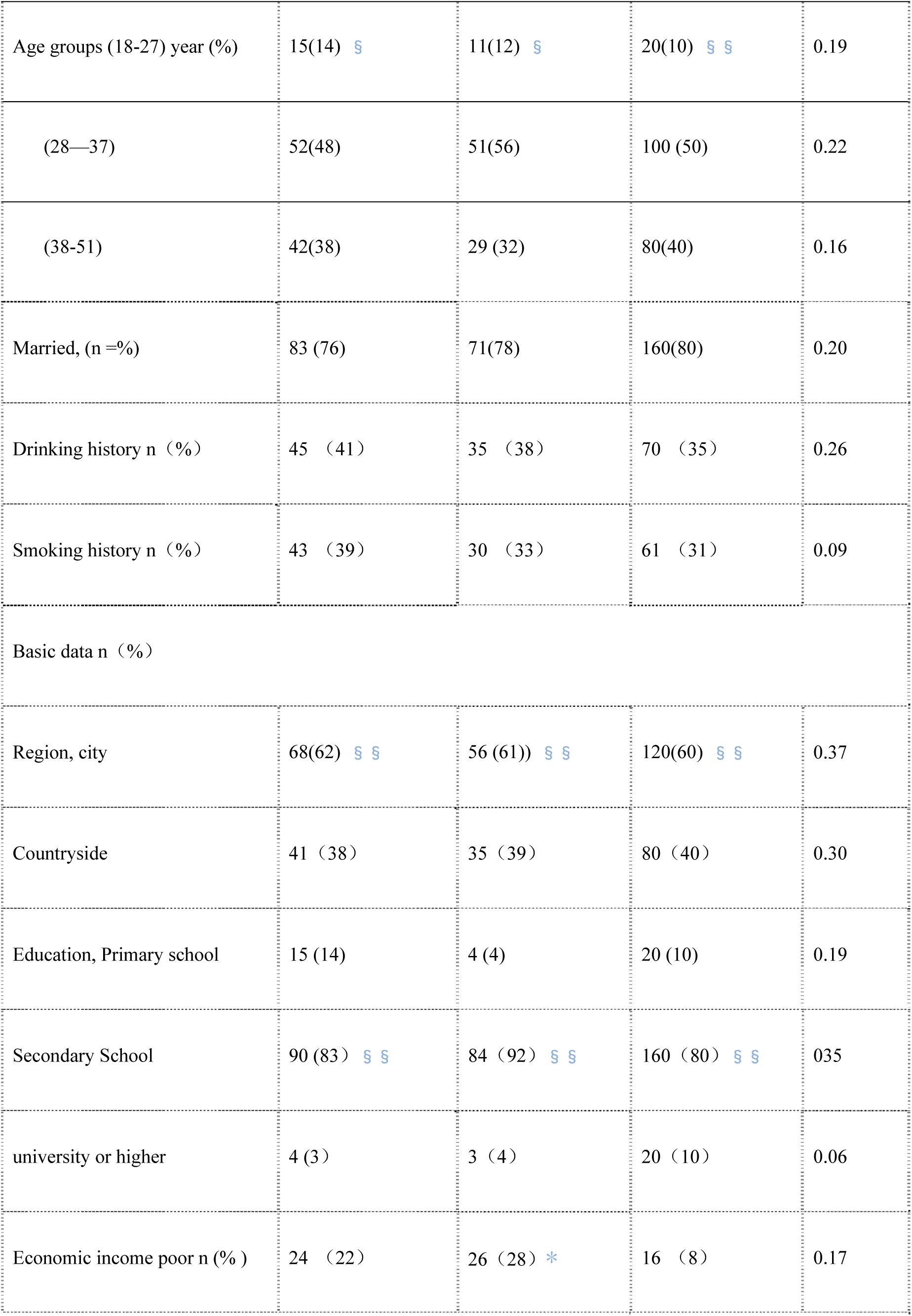
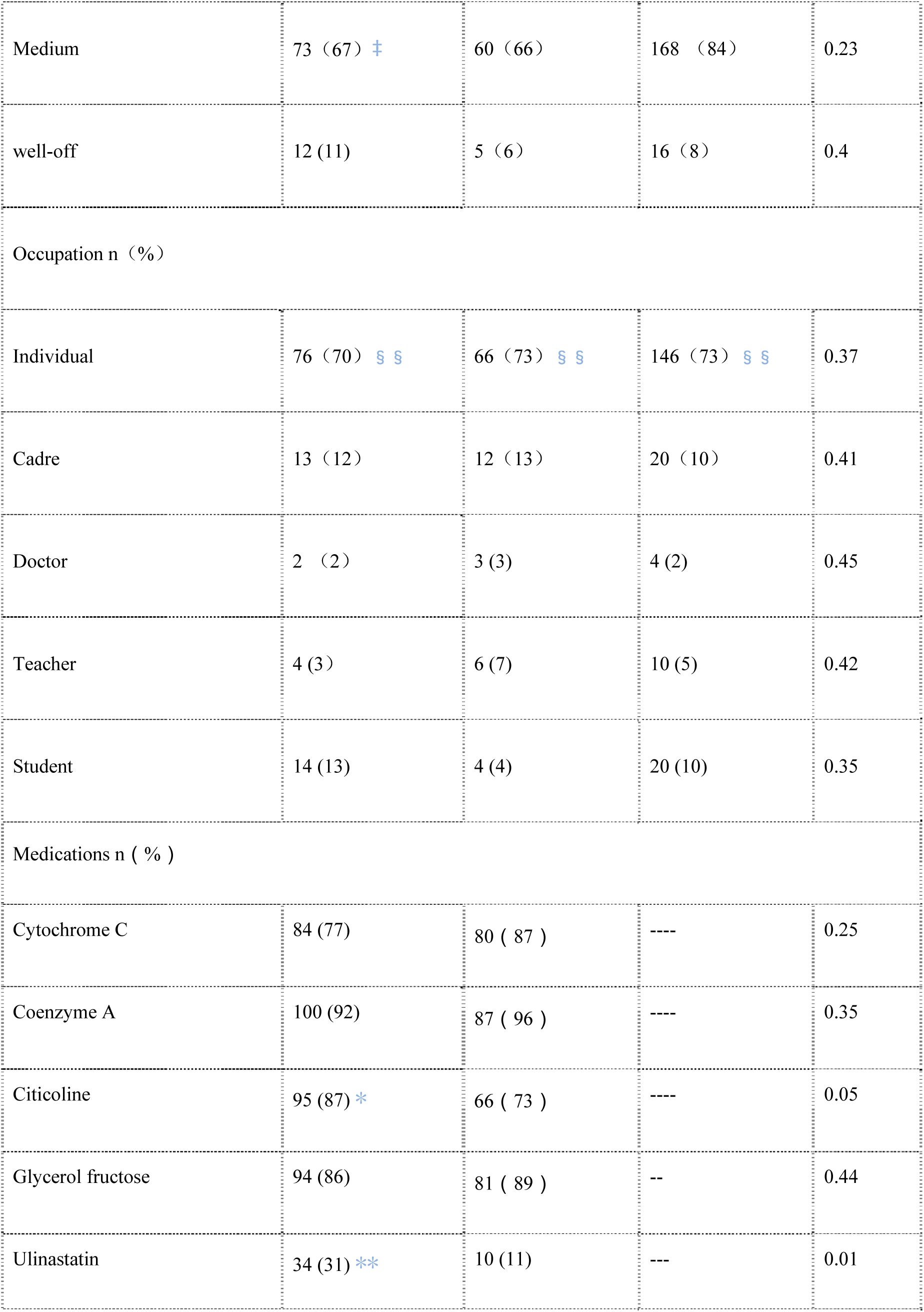
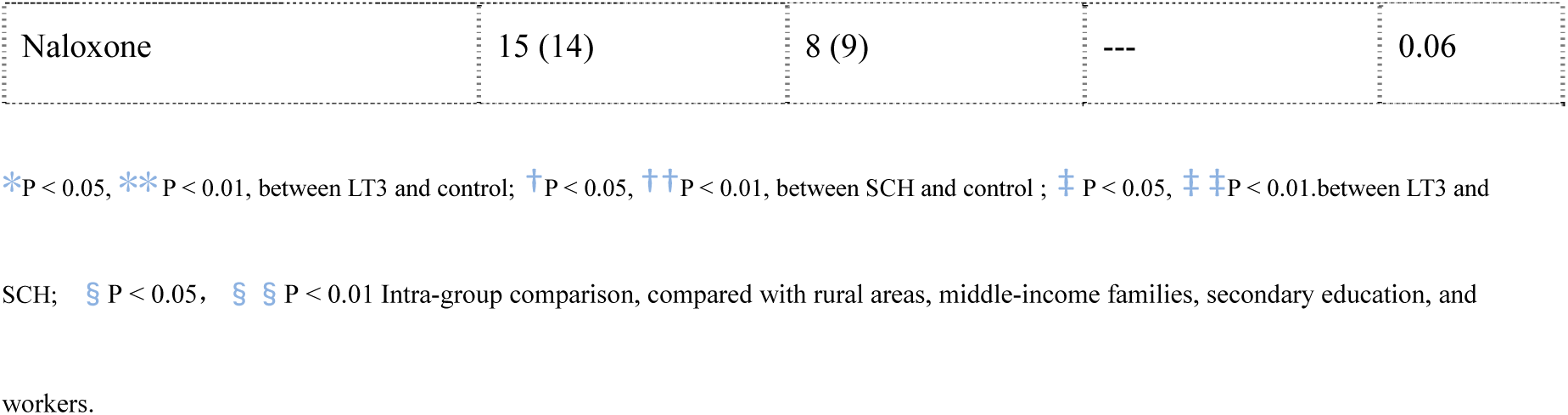
Characteristics of the study population

| variable | LT3 (n = 109) | SCH (n = 91) | Control(n=200) | P value |
| --- | --- | --- | --- | --- |
| Demographics |  |  |  |  |
| Male, (n =%) | 80 (73) * | 31 (34) | 100 (50) | 0.05 |
| Age groups (18-27) year (%) | 15(14) § | 11(12) § | 20(10) § § | 0.19 |
| (28—37) | 52(48) | 51(56) | 100 (50) | 0.22 |
| (38-51) | 42(38) | 29 (32) | 80(40) | 0.16 |
| Married, (n =%) | 83 (76) | 71(78) | 160(80) | 0.20 |
| Drinking history n (%) | 45 (41) | 35 (38) | 70 (35) | 0.26 |
| Smoking history n (%) | 43 (39) | 30 (33) | 61 (31) | 0.09 |
| Basic data n (%) |  |  |  |  |
| Region, city | 68(62) § § | 56 (61)) § § | 120(60) § § | 0.37 |
| Countryside | 41 (38) | 35 (39) | 80 (40) | 0.30 |
| Education, Primary school | 15 (14) | 4 (4) | 20 (10) | 0.19 |
| Secondary School | 90 (83) § § | 84 (92) § § | 160 (80) § § | 0.35 |
| university or higher | 4 (3) | 3 (4) | 20 (10) | 0.06 |
| Economic income poor n (%) | 24 (22) | 26 (28) * | 16 (8) | 0.17 |
| Medium | 73 (67) ‡ | 60 (66) | 168 (84) | 0.23 |
| well-off | 12 (11) | 5 (6) | 16 (8) | 0.4 |
| Occupation n (%) |  |  |  |  |
| Individual | 76 (70) § § | 66 (73) § § | 146 (73) § § | 0.37 |
| Cadre | 13 (12) | 12 (13) | 20 (10) | 0.41 |
| Doctor | 2 (2) | 3 (3) | 4 (2) | 0.45 |
| Teacher | 4 (3) | 6 (7) | 10 (5) | 0.42 |
| Student | 14 (13) | 4 (4) | 20 (10) | 0.35 |
| Medications n (%) |  |  |  |  |
| Cytochrome C | 84 (77) | 80 (87) | --- | 0.25 |
| Coenzyme A | 100 (92) | 87 (96) | --- | 0.35 |
| Citicoline | 95 (87) * | 66 (73) | --- | 0.05 |
| Glycerol fructose | 94 (86) | 81 (89) | -- | 0.44 |
| Ulinastatin | 34 (31) ** | 10 (11) | --- | 0.01 |
| Naloxone | 15 (14) | 8 (9) | --- | 0.06 |

\* $P < 0.05$ , \*\* $P < 0.01$ , between LT3 and control; † $P < 0.05$ , †† $P < 0.01$ , between SCH and control; ‡ $P < 0.05$ , ‡‡ $P < 0.01$ , between LT3 and
SCH; § $P < 0.05$ , §§ $P < 0.01$ Intra-group comparison, compared with rural areas, middle-income families, secondary education, and
workers.

**Figure 1.**
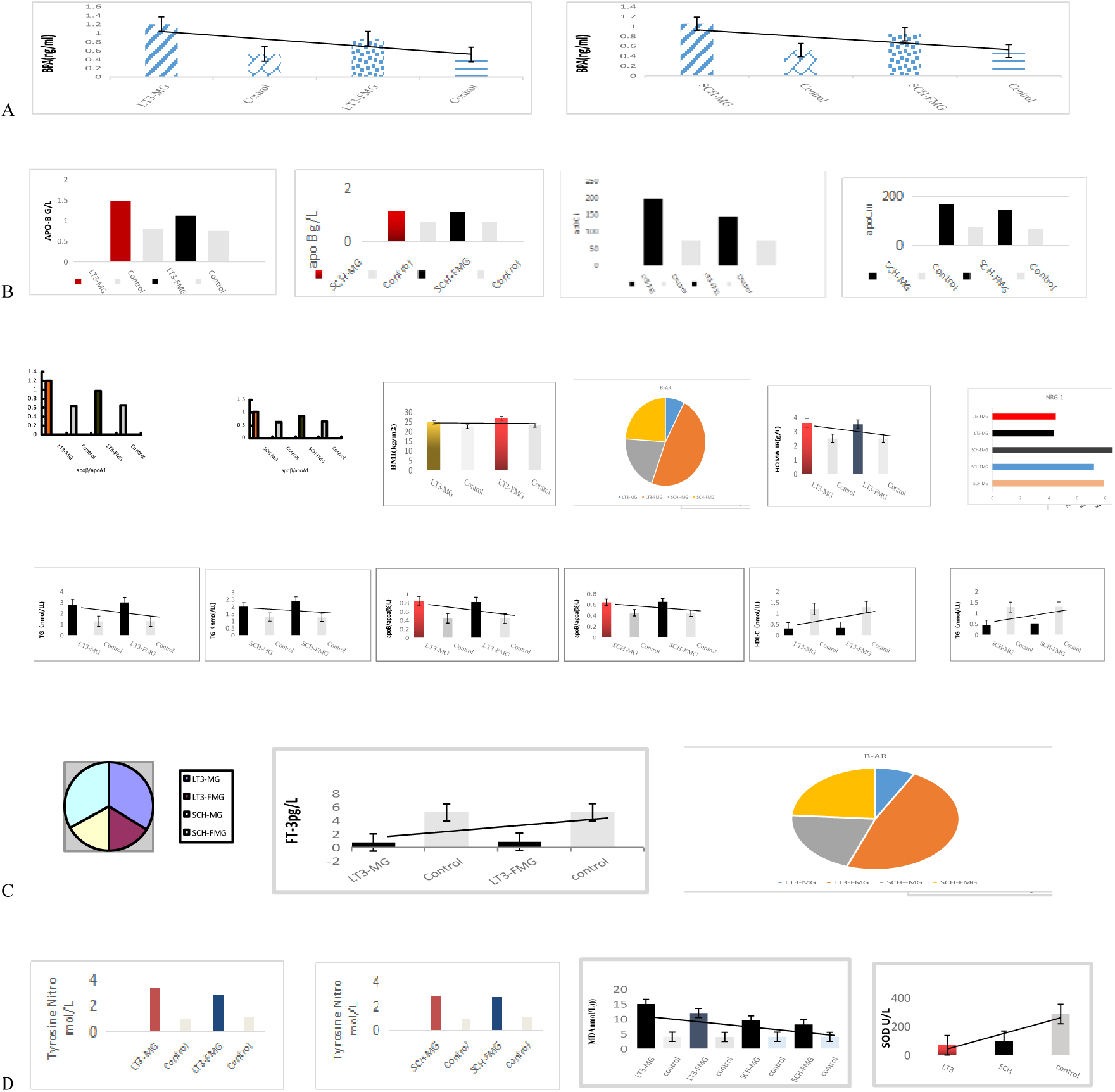
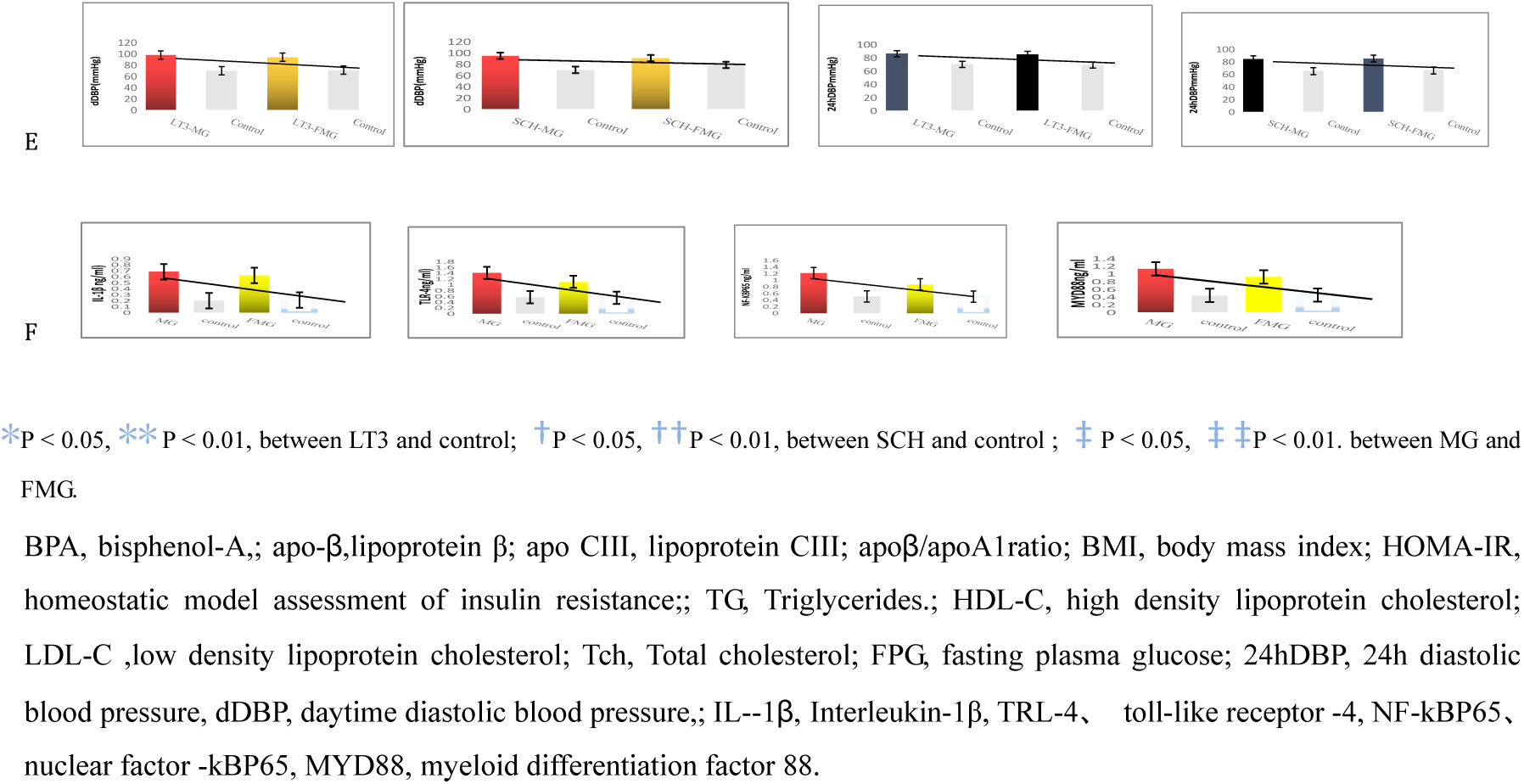
Characteristics of serum lipid metabolism and oxidative stress products in patients with low T3 and gender differences between groups

### Comparison of OS and lipid regulatory protein expression between groups

Of the 200 ICH patients,70 was obese (BMI ≥25 kg/m2) and 48 (23%) were in T3 group, including 40 females and 16 males. SCH group included 4 patients, 2 male and 2 female. Oxidative stress (OS) risk stratification; Firstly, the four main risk factors for OS (hypertension, diabetes, atherosclerosis and obesity) and secondary risk factors (smoking, hyperlipidemia, BMI≥2.5, abdominal obesity, insulin resistance and abnormal glucose tolerance) were investigated. Then, according to the direct determination of brain tissue reactive oxygen species (ROS) content: low degree 40-79, 1-2 secondary risk factors; Moderate intensity 80-119, 1 major risk factor, 2 or more minor risk factors; High intensity 120-160, more than 2 major risk factors.

FIG. 2. A: Compared with SCH group, brain tissue reactive oxygen species (ROS) in LT3 group were significantly increased, and LT3-FMG-B, MG-b subgroup were significantly increased compared with LT3-FMG-a and MG-a. B: Expression of β3-AR and UCP1 was decreased in LT3 group compared with control group and SCH group, and more significantly in male group compared with female group. C: Compared with LT3 group, NRG-1 expression in intestinal and brain tissues of SCH group was more significantly increased;

**Figure 2.**
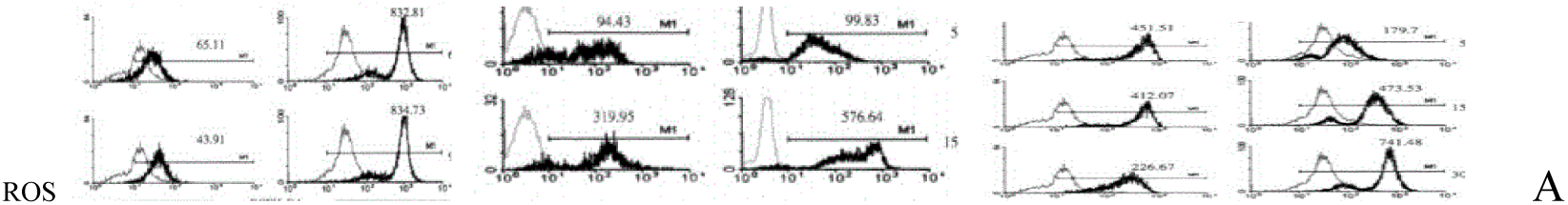
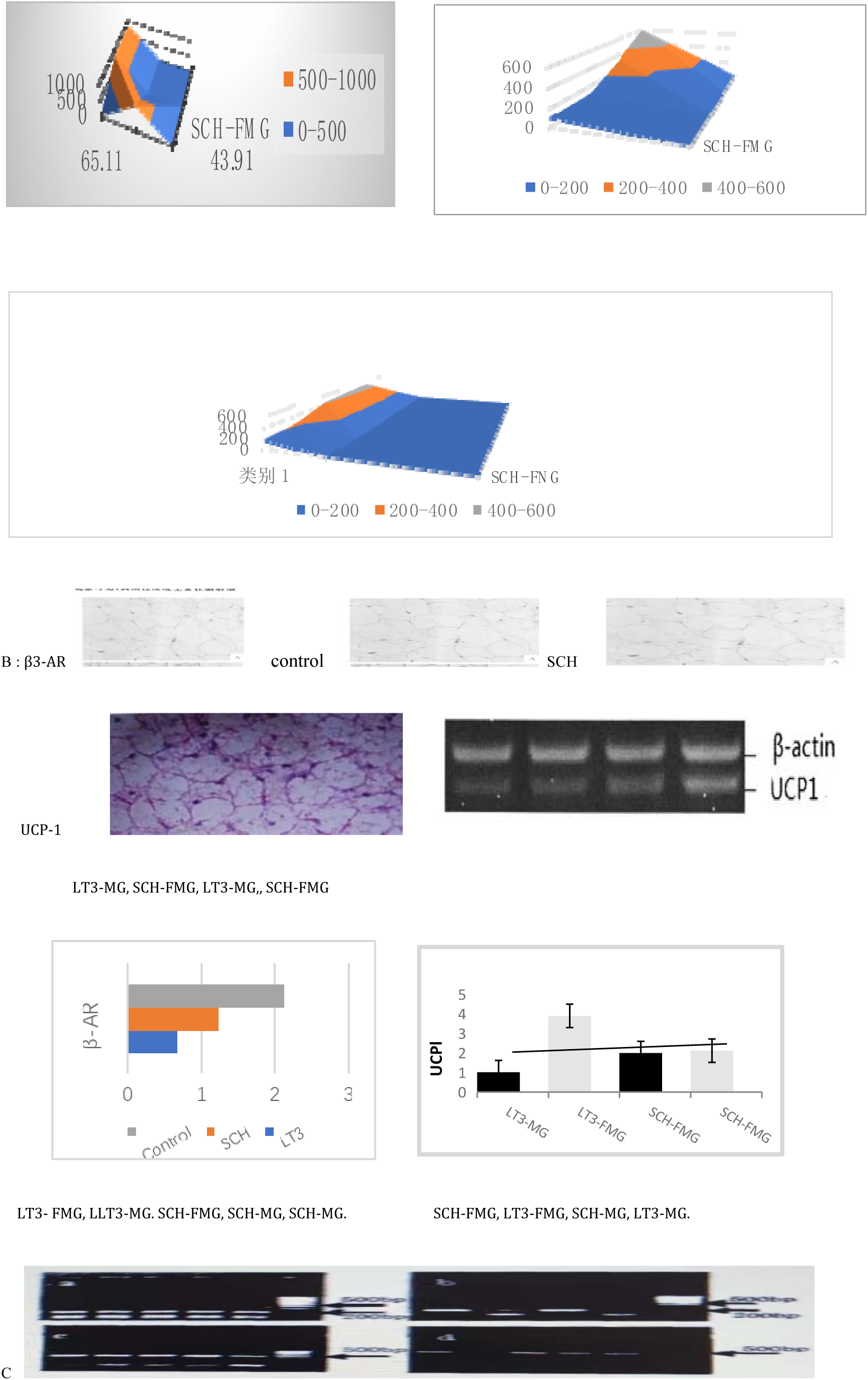
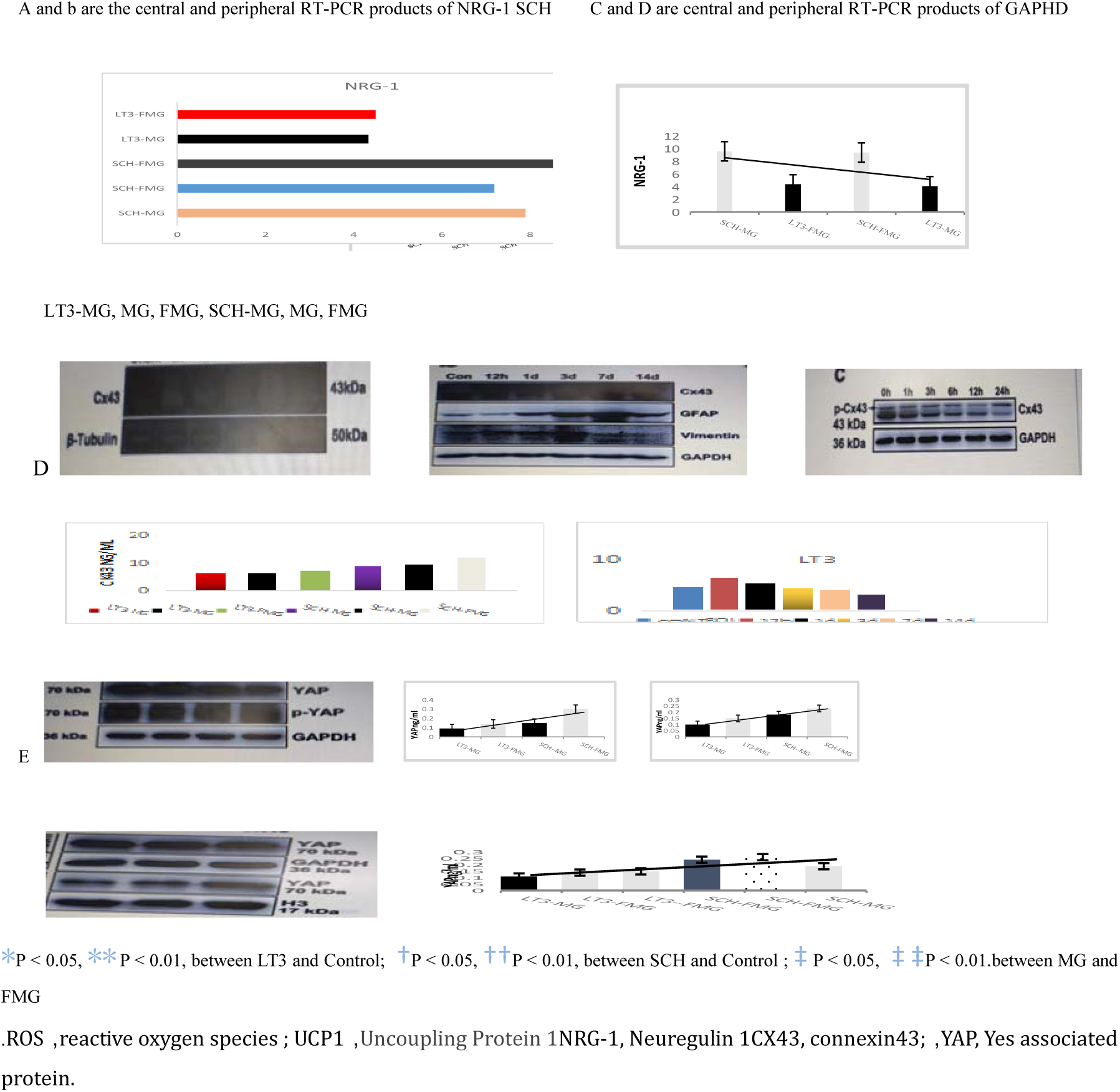
Comparison of the expression of reactive oxygen species and lipid oxidative stress messenger proteins between groups.

Compared with MG group, NRG-1 in FMG group showed an increasing trend. D: Compared with LT3 group, CX43 expression increased more significantly in SCH group. However, CX43 expression showed a temporal difference, and the expression level decreased with the increase of time. E: Compared with SCH group, the expression of LT3 group was significantly reduced.

### Basic analysis of sequencing data

The sequencing data were collected from 28 LT3 and SCH patients in ICH group and 29 stool samples in control group at baseline, thus a total of 85 stool samples were collected in this study. 7321 475 original sequences were obtained, and the number of sequences matched to a single sample ranged from 48302 to 109837, with an average of 86 135 sequences per sample. The processed sequence results are obtained by reading, assembling and quality control of the original data. After chimeric filtering, valid data can be obtained for subsequent analysis. The valid data was 6 361 825, with an average of 74 845 per sample, and the actual range was 39 904 –– 97 910. The average reading length was 253 bases and the sequencing depth coverage was 99.75%.

The degree of intra-group species diversity was determined in three groups in this study: SCH group, LT3 group and control group, and the alpha diversity index between the three groups was analyzed. Except for goods-coverage, there were significant differences in the diversity of alpha between LT3 group and control group. Compared with the control group, the alpha indexes of SCH group showed no statistical significance except shannon index. The alpha diversity index of LT3 group was not statistically significant compared with SCH group. The composition of different groups of microbiome, Principle coordinates analysis (PCoA) (Table 2, figure 3 See supplementary data online)

**Figure 3.**
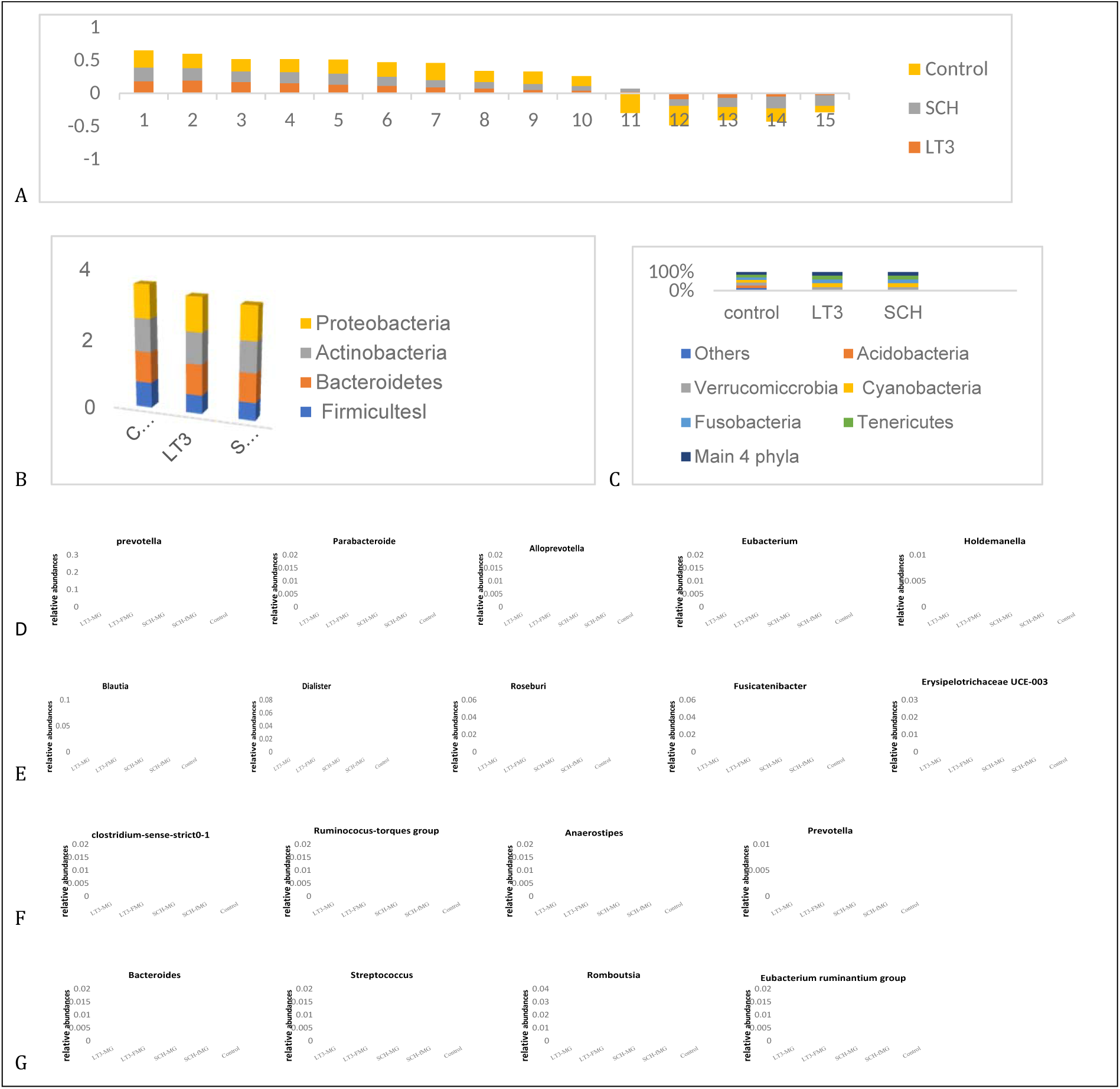
Comparison of intestinal flora between groups

Figure 4, A: Pearson correlation analysis was used to analyze the relationship between lipid analysis, serum OS and intestinal flora, and correlation analysis was conducted with the relative abundance of each flora. For ICH, body mass change is the main indicator of concern. ICH was correlated with bifidobacterium abundance, and the correlation coefficient was –0,51. The relative abundance of Clostridium was positively correlated with the variation value of OS parameter, and the correlation coefficient was 0.48. There was a correlation between Bacteroides and ROS values, and the correlation coefficient was –0.55, all (P < 0.01).

**Figure 4.**
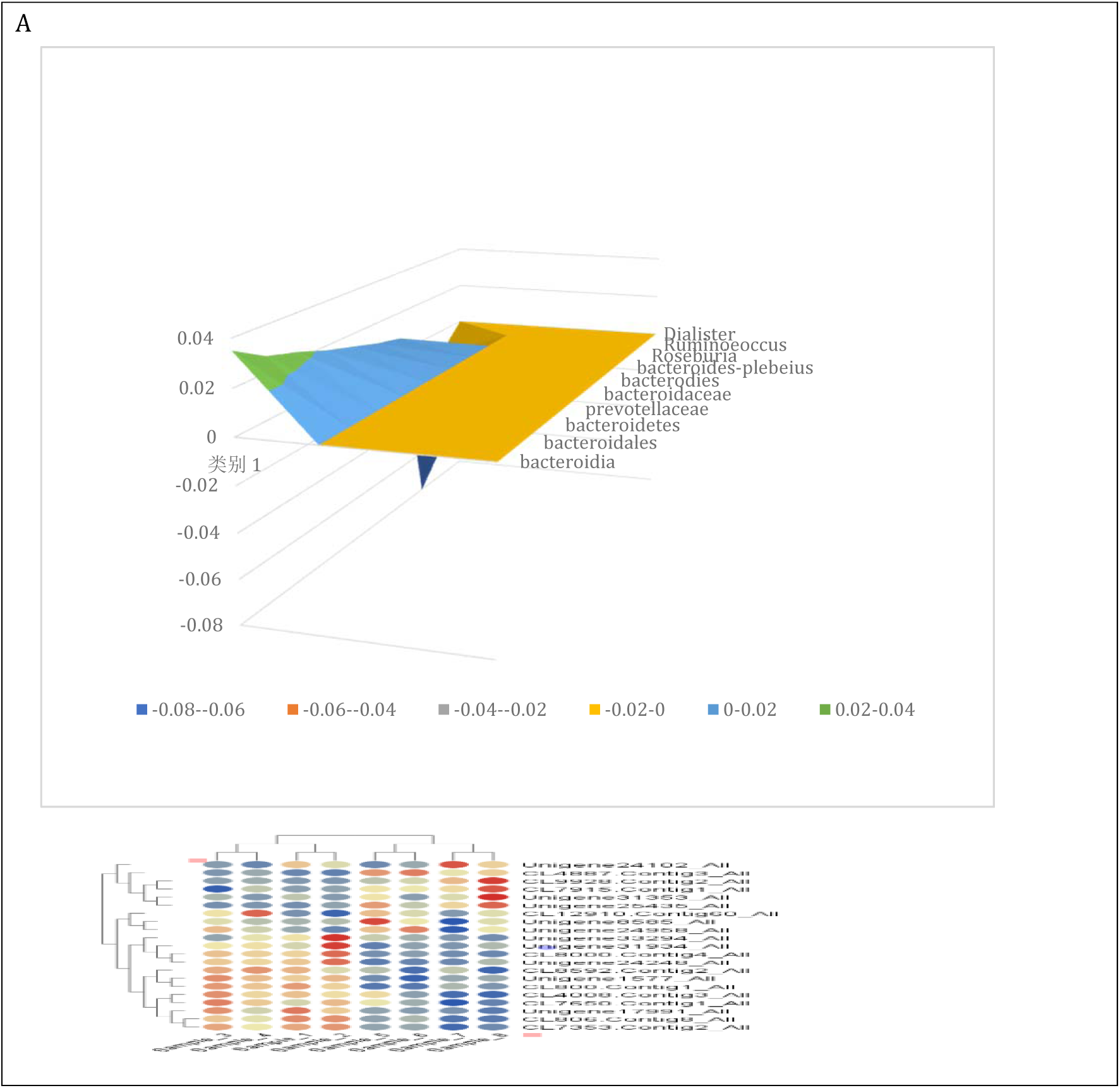
Linear discriminant analysis effect size analysis between LT3 and control

Figure 5A: Serum glial fibrillary acidic protein (GFAP) was measured using enzyme-linked immunosorbent assay (Elisa) double antibody sandwich. Compared with the control group, there was a significant increase in GFAP in the LT3 group compared with the SCH group, while there was also a statistically significant difference in the increase in GFAP in the LT3a group compared with the SCH group. B: Serum anti-TMEM119 antibody was determined by immunohistochemical staining and immunofluorescence. The expression of TMEM in the LT3 and SCH groups was reduced compared with the control group. C: Compared with SCH group, LT3 group showed significantly increased expression of astrocytes and oligodendrocyte progenitor cells in direct contact.

**Figure 5.**
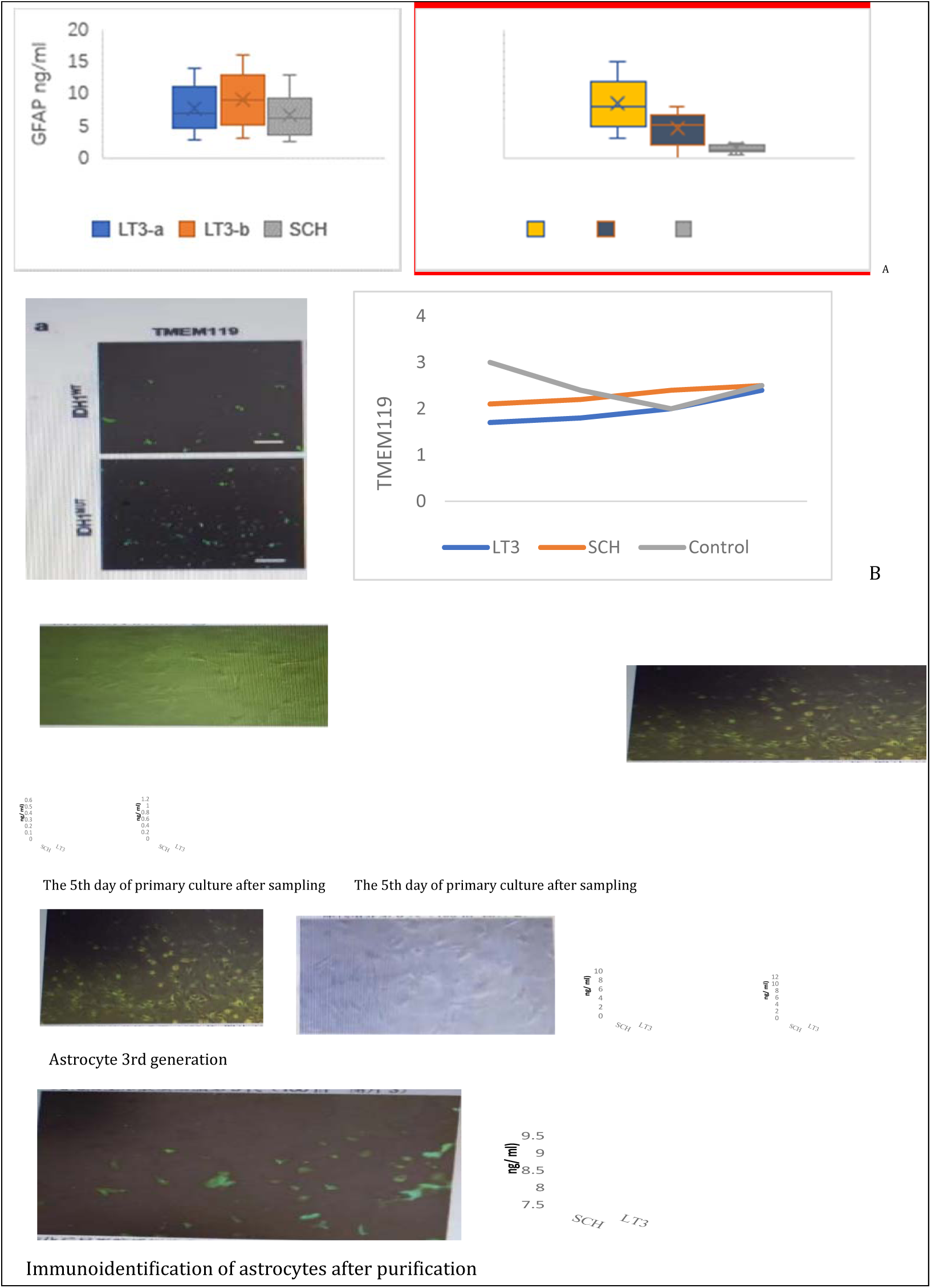
Comparison of astrocyte and serum glial fibrillary acid protein and anti-TMEM119 antibody expression groups

FIG 6 A, B and C are LT3 group, respectively. The significant relationship between SCH group and Control is generally shown in the heat map. D and E are Pearson correlation heat maps of differences in serum lipid metabolism and oxidative stress parameters and alternate flora abundance between LT3 and SCH groups.

**Figure 6.**
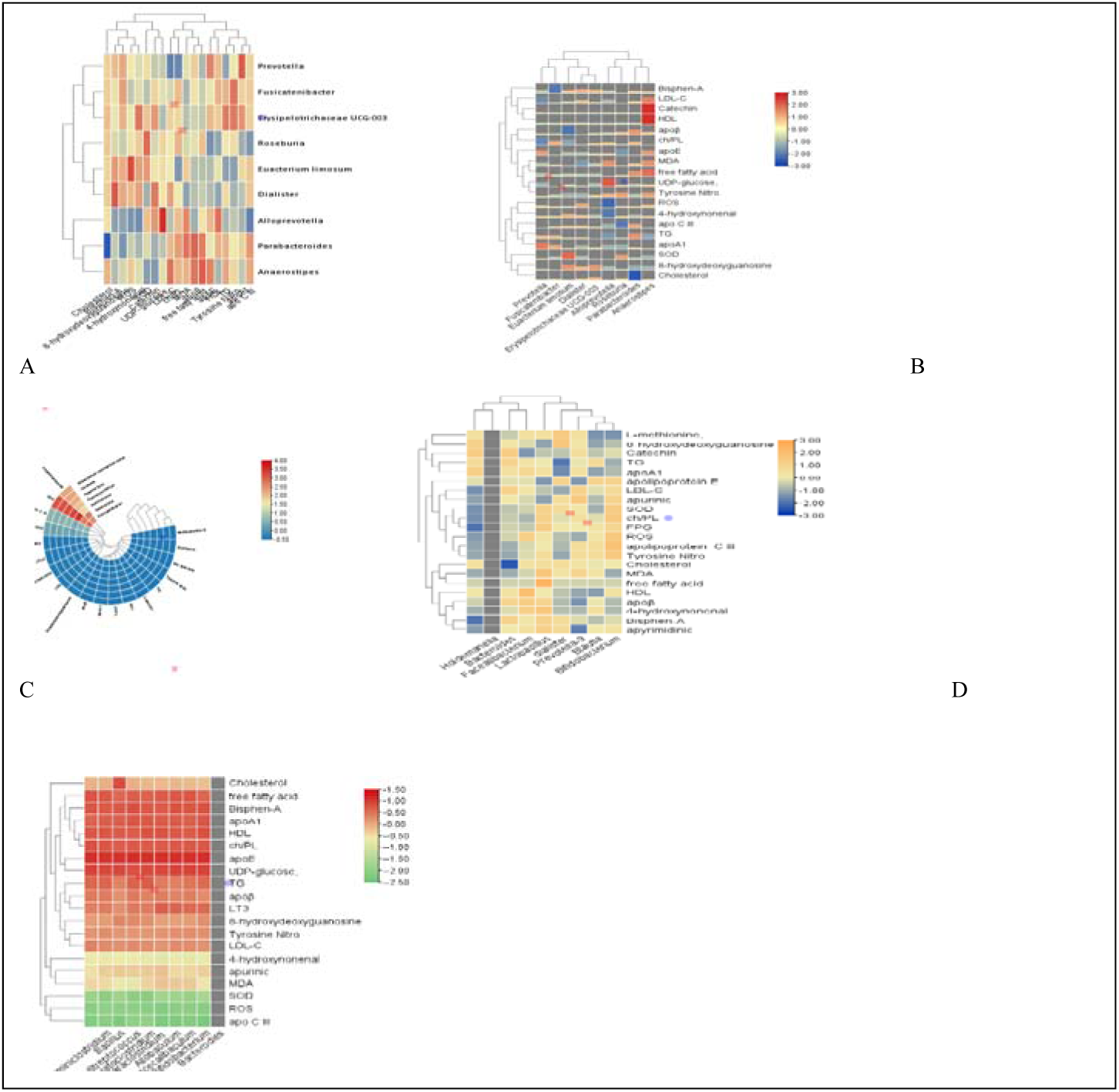
Heat maps for clustering analysis of lipid metabolites and flora abundance in ICH and control groups, LT3 and SCH groups

### Relationship between BPA and risk factors of OS expression

The BPA and ROS curve was drawn. The area under the curve was 0.744 (95%CI 0.624-0.894), the maximum Uden index was 0.426, the sensitivity was 75.4%, the specificity was 67.4%, and the corresponding critical value was 1.567. After adjusting for confounding factors such as gender, age and marital status, the risk of serum BPA and MDA, SDA in ICH patients were 5.12 and 6.14 times that of the general population.

## DISCUSSION

Bisphenol A(BPA), as a heterogeneous estrogen, does not directly affect plasma lipid levels, but enhances palmitoacylation of differentiated cluster 36 (CD36) expression in macrophages and is associated with lipid accumulation ^17, 18^. Produce low inflammatory state, induce liver toxicity and intestinal ecological imbalance, reduce superoxide dismutase (SOD), glutathione (GSH), increase malondialdehyde (MDA) level, increase oxidative stress (OS) level ^9^. Neurotoxic effects and endocrine dysfunction, showing that neuronal structure and Neuregulin –1 (NRG-1) in other parts of the intestinal wall exert a moderated protective effect on nerve cells 11. Furthermore, OS may be a key driver of ICH development, contributing to the identification of stress-related functional changes and disease phenotypes, and the OS response may increase neuronal dysfunction and immune cell adhesion ^19, 20^. β3-adrenergic receptors (β3AR) integrate thyroid hormone signals for adipocyte thermogenesis and lipogenesis, and the central nervous system’s ability to regulate energy balance and metabolism ^10, 21^. It can also reduce the release of myeloid monocytes after stroke, promote the stress effect of circulating alarm protein and reduce the activation of systemic monocytes and major artery invasion^22^, NRG-1 maintains the integrity of the blood-brain barrier and reduces the OS response. Our study showed that compared with the control group, patients with bisphenol A-related ICH presented lipid metabolism disorders, increased apoβ / apoA1 ratio, decreased serum SOD and β3-AR expression, and increased serum MDA, brain reactive oxygen species (ROS) and NRG-1 levels. Three sex differences were also found: (1) Compared with female patients (FMG), serum BPA, apoβ and apo CIII(apo CIII) increased in male patients (MG), and the increase was more significant in LT3 group; (2) Compared with FMG, the increase of 3-nitrotyrosine was more significant in MG patients, as well as in LT3 group; (3) Compared with MG, the increase of body mass index (BMI) in FMG was more obvious. These results indicate that specific environmental risk factors lead to significant gender differences in lipid metabolism and oxidative stress response in the body, and the two respond to each other and jointly promote such a pathophysiological super reaction process.

Ecological disorders (intestinal bacterial imbalance) are associated with metabolic syndrome (MetS) and estrogen homeostasis. Intestinal flora can affect the human lipid metabolic stress system through the bacteria itself or its metabolites, and the influence can also activate the immune system and nervous system through blood circulation. Therefore, the measurement of serum lipids and OS can also reflect the differences of intestinal microorganisms. In ICH patients, Prevotella, Parabacteroides, isoprevotella, Eubacteria mucoides and Haldemanella had higher abundance than controls. The abundance of Eubacterium myxus, Microbacillus, Roxella, Firmicutes and Streptobacillus spindus decreased. LEFSe analysis showed that Prevotella and Bacteroides were enriched, while Eubacillus myxus, Microbacillus, and Rhodotella were enriched in the control group. Previous studies on ICH have reported that ^14^, in the genus level streptococcus, bifidobacterium, Akermannia mucophilus and Lactobacillus are more abundant. In another report of hypertensive intracerebral hemorrhage ^23^, streptococcus and mucous eubacteria were associated with increased blood pressure. The differences among these different studies may be related to etiology and heterogeneity. Our clinical study found that in the correlation analysis characterized by lipid metabolism and oxidative stress products, ROS was positively correlated with the abundance of bifidobacterium and Bacteroides in ICH patients with a correlation coefficient of 0,51,0.52, and LT3 was negatively correlated with the relative abundance of Clostridium difficile with a correlation coefficient of –0.48, all (P < 0.01).

Messenger connexin (CX43) regulates antioxidant stress response^4^, and YAP is a transcriptional coactivator. Down-regulation of Cx43 induces YAP nuclear translocation, which helps to control astrocyte activation and drive microglia activity ^25^. The integrity of the blood-brain barrier is also consistent with astrocyte function ^26^, will help regulate glutamate and ion homeostasis, cholesterol and sphingolipid metabolism, calcium kinetics, glial cell transport, metabolism, and inflammation, and respond accordingly to environmental factors ^27^. Oxidative stress and mitochondrial dysfunction are involved in astrocyte homeostasis. Astrocyte activation and promotion of inflammatory activity, causing YAP signaling to form reactive astrocyte disturbances ^28^. In this study, TLR4-NF-κB signaling pathway series factors increased significantly, and initial CX43 concentration and changes in CX-43/YAP axis may be associated with higher risk after ICH. It is speculated that this phenomenon is related to treatment and the protective effect of autoimmune.

In conclusion, BPA can induce vasotoxic risk events, gradually acting on cerebrovascular pathology and molecular phenotypic biochemical factors, and inflammatory neuromicroenvironment may be associated with neuronal apoptosis. Oxidative stress induced by the LTRs-NFκBp65 inflammatory pathway is associated with different subtypes or phenotypes of immune cells and activation of the inflammatory cascade after stroke events.^28^ Cx43/YAP cross-talk induces reactive astrocyte hyperplasia in ICH. The importance of Cx43 in anti-oxidative stress lies in the release of functional mitochondria as part of the homeostasis regulation process of astrocytes.

## Conclusion

BPA can induce vasotoxic risk events, gradually acting on cerebrovascular pathology and molecular phenotypic biochemical factors, and inflammatory neuro-microenvironment may be associated with neuronal apoptosis. Oxidative stress induced by the LTRs-NFκBp65 inflammatory pathway is associated with different subtypes or phenotypes of immune cells and activation of the inflammatory cascade after stroke events. Cx43/YAP axis induces reactive astrocyte hyperplasia in ICH. The importance of Cx43 in anti-oxidative stress lies in the release of functional mitochondria as part of the homeostasis regulation process of astrocytes.

## Data Availability

I share the responsibility with other members of the project team. All authors agree and support the submission of this manuscript to your journal for publication. In the long-term clinical practice, encounter a large number of young patients with cerebral hemorrhage can not find the cause of bleeding, for unexplained cerebral hemorrhage. These patients had no history of hypertension, and no brain tumors or vascular malformations were detected by CT and magnetic resonance imaging. Therefore, I first made a preliminary experiment to examine the serum bisphenol A level in one of the patients. The results showed that BPA increased significantly in this patient. Therefore, the subject has been declared.No, I worked with other authors. They have doctoral supervisors, chief physicians and professors. I am a professor and chief physician of cardiovascular medicine.

## Nonstandard Abbreviations and Acronyms

BPA: bisphenol
ICH: intracerebral hemorrhage
LTs3: low T3 Syndrome
OS: oxidative stress
IFD: Intestinal flora dysregulation
GBA: gut-brain axis

## ACKNOWLEDGEMENTS

Thanks to Professor Wu Bowei, Zhang Mingshen and some teachers, residents, students of Shanxi Medical University.

## References

1. Johnson CO, Nguyen M, Roth GA, Nichols E, Alam T, Abate D. Global regional, and national burden of stroke, 1990-2016: a systematic analysis for the Global Burden of Disease Study 2016. Lancet Neurol. 2019; 18:439–458. doi: 10.1016/S1474-4422(19)30034-1.

2. Valery L. Feigin, Grant Nguyen, & Kelly Cercy, B.S., Global, Regional, and Country-Specific Lifetime Risks of Stroke, 1990 and 2016. The GBD 2016 Lifetime Risk of Stroke Collaborators. N Engl J Med. 2018; 379:2429–2437. doi. 10.1056/NEJMoa1804492.

3. Virani SS, Alonso A, Benjamin EJ, Bittencourt MS, Callaway CW, Carson AP, et al. Heart disease and stroke statistics-2020 update:a report from the American Heart Association. Circulation. 2020;141: e139–e596.

4. Xiao Chen, Huaibin Liang, Zhiyu Xi, Yong Yang, Huimin Shan, Baofeng Wang, Zhihong Zhong. BM-MSC Transplantation Alleviates Intracerebral Hemorrhage-Induced Brain Injury, Promotes Astrocytes Vimentin Expression, and Enhances Astrocytes Antioxidation via the Cx43/Nrf2/HO-1 Axis. Front Cell Dev Biol. 2020; 8: 302. doi: 10.3389/fcell.2020.00302.

5. Samuel X. Shil, Kaibin Shil, Qiang Liu. Brain injury instructs bone marrow cellular lineage destination to reduce neuroinflammation. Science Translational Medicine.2021;13 (589): eabc7029.

6. Abel Eraso-Pichot, Sandrine Pouvreau, Alexandre Olivera-Pinto, Paula Gomez-Sotres, Urszula Skupio, Giovanni Marsicano. Endocannabinoid signaling in astrocytes. Glia. 2023; 71: 44–59. doi: 10.1002/glia.24246

7. Francesca Farrugia, Alexia Aquilina, Josanne Vassallo, Nikolai Paul Pace. Bisphenol A and Type 2 Diabetes Mellitus. Int J Environ Res Public Health. 2021;18:716–722. doi:10.3390/ijerph18020716.

8. Ya Maa Haohao, Liua Jinxia, Wua Le, Yuana Yueqin, Wanga Xingde, Dua Rui Wang. The adverse health effects of bisphen A and related toxicity mechanisms. Environmental Research. 2019; 10:8575–8581. doi.org/10.1016/j.envres.

9 Ruijing Liu, Boping Liu, Lingmin Tian, Xinwei Jiang, Xusheng Li, Dongbao Cai, Jianxia Sun, Weibin Bai, Yulong Jin. Exposure to Bisphenol A Caused Hepatoxicity and Intestinal Flora Disorder in Rats. Int J Mol Sci. 2022 Jul; 23(14): 8042. doi: 10.3390/ijms23148042.

10 Adilson Guilherme, Batuhan Yenilmez, Alexander H. Bedard, Felipe Henriques, Dianxin Liu, Alexandra Lee, Lauren Goldstein, Mark Kelly, Sarah M. Nicoloro, Min Chen, et al Adipocyte Thermogenesis and Lipogenesis through γ3-Adrenergic and Thyroid Hormone Signal Integration. Cell Rep. 2020;31: 107598.doi: 10.1016/j.celrep.2020.107598.

11 Kamila Szymańska, Krystyna Makowska, Jarosław Całka, Sławomir Gonkowski. The Endocrine Disruptor Bisphenol A Affects the Enteric Neurons Immunoreactive to Neuregulin 1 (NRG1) in the Enteric Nervous System of the Porcine Large Intestine. Int J Mol Sci.2020;21: 8743–8749. doi: 10.3390/ijms21228743,

12 Warner, G.R., Flaws, J. A. Bisphenol-A and phthalates: How environmental chemicals are reshaping toxicology. Toxicol Sci.2018; 166:246–249.

13. Wang Yupeng, Zhang Ye Li, Xiangbin Zhang, Junyi Ma, Xuelun Zou, TianXing Yao, Si Li, Junyou Chen, Huifang Zhou, Lianxu Wu, et al. Multi-omics reveals specific host metabolism-microbiome associations in intracerebral hemorrhage Front Cell Infect Microbiol. 2022; 12: 999627. Dec 22. doi: 10.3389/fcimb.2022.999627

14. WHO Expert consultation. Appropriate body-mass index for Asian populations and its implications for policy and intervention strategies Lancet. 2004;363:157-163. doi: 10.1016/SO140-6736(03)15268-3.

15. J Claude Hemphill III, Steven M Greenerg, Craig S Anderson, Guideline for the Management of Spontaneous Intracerebral Hemorrhage. Stroke.2015;46:2032–2060.

16. 1999 World Health Organization-International Society of Hypertension Guidelines for the Management of hypertension. Clinical and Experiment Hypertension. 2009; 21:1009–1060 151-183.

17. Katarzyna Szkudelska, Monika Okulicz, Tomasz Szkudelski. Bisphenol A disturbs metabolism of primary rat adipocytes without affecting adipokine secretion. Environ Sci Pollut Res Int. 2021; 28:23301–23309.doi: 10.1007/s11356-021-12411-0.

18 Yun Zhang, Doudou Dong, Xiaoting Xu, Hui He, Yu an Zhu, Tingwen Lei, Hailong Ou. Oxidized high-density lipoprotein promotes CD36 palmitoylation and increases lipid uptake in macrophages. J Biol Chem. 2022 Jun; 298(6): 102000.doi: 10.1016/j.jbc.2022.102000.

19. Jun Bai, Renbo Tan, Zheng An, Ying Xu. Quantitative estimation of intracellular oxidative stress in human tissues. Brief Bio-inform. 2022; 23:206–214. doi: 10.1093/bib/bbac206.

20. Tracy D. Chung, Raleigh M. Linville, Zhaobin Guo, Robert Ye, Ria Jha, Gabrielle N. Grifno, Peter C. Effects of acute and chronic oxidative stress on the blood–brain barrier in 2D and 3D in vitro models, Searson Fluids Barriers CNS. 2022; 19: 33–41. doi: 10. 1186/ s12987-022-00327-x

21. Jennifer E. Richard, Lorena López-Ferreras, Belén Chanclón, Kim Eerola, Peter Micallef, Karolina P. Skibicka, Ingrid Wernstedt Asterholm. CNS β3– adrenergic receptor activation regulates feeding behavior, white fat browning, and body weight. American Journal of Physiology-Endocrinology and Metabolism. 2017;313:32017. doi.org/10.1152/ajpendo.00418.2016.

22. Stefan Roth, Vikramjeet Singh, Steffen Tiedt, Lisa Schindler, Georg Huber, Arie Geerlof, Daniel J. Antoine, Antoine Anfray, Cyrille Orset, Maxime Gauberti, et al. Brain-released alarmins and stress response synergize in accelerating atherosclerosis progression after stroke,Science Translational Medicine. 2018;13: 3–14, doi: 10.1126/scitranslmed.aao1313.

23. Dong Yan, Ye Sun, Xiaoyue Zhou, Wenhao Si, Jieyu Liu, Min Li, Minna Wu. Regulatory effect of gut microbes on blood pressure. Animal Model Exp Med. 2022; 5: 513–531. doi: 10.1002/ame2.12233.

24. Yong Yang, Jie Ren, Yuhao Sun, Yuan Xue, Zhijian Zhang, Aihua Gong, Baofeng Wang, Zhihong Zhong, Zhenwen Cui, Zhiyu Xi, et al. connexin43/YAP axis regulates astroglial-mesenchymal transition in hemoglobin induced astrocyte activation. Brain Cell Death Differ. 2018; 25: 1870–1884. doi: 10.1038/s41418-018-0137-0.

25 Yan Zhang, Suliman Khan, Yang Liu, Rabeea Siddique, Ruiyi Zhang, Voon Wee Yong, Mengzhou Xue. Gap Junctions and Hemichannels Composed of Connexins and Pannexins Mediate the Secondary Brain Injury Following Intracerebral Hemorrhage. Biology (Basel) 2022; 11: 27–34. doi: 10.3390/biology11010027.

26. Hailong Yu, Xiang Cao, Wei Li, Pinyi Liu, Yuanyuan Zhao, Lilong Song, Jian Chen, Beilei Chen, Wenkui Yu, Yun Xu..Targeting connexin 43 provides anti-inflammatory effects after intracerebral hemorrhage injury by regulating YAP signaling. J Neuroinflammation. 2020; 17: 322-331. doi: 10.1186/s12974-020-01978-z

27. Nicholas W. Kieran, Rahul Suresh, Marie-France Dorion, Adam MacDonald, Manon Blain, Dingke Wen, Shih-Chieh Fuh, Fari Ryan, Roberto J. Diaz, Jo Anne Stratton, et al. MicroRNA-210 regulates the metabolic and inflammatory status of primary human astrocytes. J Neuroinflammation. 2022; 19: 10–18. doi: 10.1186/s12974-021-02373-y

28. Rezan Ashayeri Ahmadabad, Zahra Mirzaasgari, Ali Gorji, Maryam Khaleghi Ghadiri. Toll-Like Receptor Signaling Pathways: Novel Therapeutic Targets for Cerebrovascular Disorders. Int J Mol Sci. 2021;22: 6153. doi: 10.3390/ijms22116153.

29 Xiaowei Fei, Yeting He, Jia Chen, Weitao Man, Chen Chen, Kai Sun, Boyun Ding, Chongwu Wang, Ruxiang Xu. The role of Toll-like receptor 4 in apoptosis of brain tissue after induction of intracerebral hemorrhage. Journal of Neuroinflammation. 2019 16:234–243.

